# Characterization of patients that can continue conservative treatment for adenomyosis

**DOI:** 10.1101/2021.03.03.21252870

**Authors:** Chiho Miyagawa, Kosuke Murakami, Takako Tobiume, Takafumi Nonogaki, Noriomi Matsumura

**Affiliations:** Department of Obstetrics and Gynecology, Kindai University Faculty of Medicine, Osaka-sayama, Osaka, Japan; Department of Obstetrics and Gynecology, Osaka Red Cross Hospital, Osaka, Japan; Department of Obstetrics and Gynecology, National Hospital Organization Osaka National Hospital, Osaka, Japan

## Abstract

**Introduction:** Historically, hysterectomy has been the radical treatment for adenomyosis. However, some patients do not wish to hysterectomy. Nevertheless, patients often required hysterectomy during the course of conservative treatment, but the factors involved remain unknown. The purpose of this study was to determine which patients can continue conservative treatment for adenomyosis.

**Materials and Methods:** We selected women diagnosed with adenomyosis and provided with conservative treatment at the Kindai University Hospital and Osaka Red Cross Hospital. Age at diagnosis, parity, uterine size, subtype of adenomyosis, type of conservative treatment, and timing of hysterectomy for cases with difficulty continuing conservative treatment were examined retrospectively.

**Results:** A total of 885 patients were diagnosed with adenomyosis, and 124 started conservative treatment. Conservative treatment was continued in 96 patients (77.4%) and hysterectomy was required in 28 patients (22.6%). The cumulative hysterectomy rate was 32.4%, and all women had hysterectomy within 63 months. In the decision tree analysis, 82% (23/28) of women aged 46 years or younger were able to continue conservative treatment when parity was zero or one. If parity was two and over, 95% (20/21) of those aged 39 years and older had hysterectomy.

**Conclusions:** Patients that continue conservative treatment for approximately 5 years are more likely to have successful preservation of the uterus. Multipara and higher age of diagnosis are factors that contribute to hysterectomy after conservative treatment. Parity and age at diagnosis may be stratifying factor in future clinical trials on hormone therapy.

## 1. Introduction

Adenomyosis is a benign disorder in which the endometrium and endometrial stromal cells proliferate in the muscle layer of the uterus.^1 2^ Associated symptoms of anemia, abdominal pain, and chronic pelvic pain due to excessive menstruation and dysmenorrhea are common in women of reproductive age and significantly impair quality of life.^3^ Traditionally, adenomyosis was often first diagnosed by pathological examination after hysterectomy and was considered a disorder that affected the peri-menopausal period.^4 5^ However, with the widespread use of ultrasonography and magnetic resonance imaging (MRI) in recent years, it has become possible to accurately diagnose adenomyosis by imaging, and it is now diagnosed in relatively young women.^6-9^

Historically, the radical treatment for adenomyosis has been hysterectomy.^7^ However, conservative treatments of adenomyosis, such as hormone therapy and adenomyomectomy, are preferred by patients who are young and wish to preserve fertility, or do not want hysterectomy or may be at high risk for perioperative complications.^10^ For women who do not wish to become pregnant immediately, conservative treatment mainly involves hormone therapy, which is continued until menopause.^11^ However, even with hormone therapy for adenomyosis, patients often experience persistent symptoms, including pain and drug side effects, such as irregular bleeding or osteoporosis, that result in the need for hysterectomy.^10 12-14^ To date, it has been unclear which women can continue conservative treatment for adenomyosis. The identification of factors related to the success or failure of conservative treatment would greatly contribute to the choice of treatment strategy, and significantly benefit the quality of life of women and the health care economy.

The purpose of this study was to evaluate the treatment course of patients with adenomyosis who have requested conservative treatment, and to determine which women can continue conservative treatment.

## 2. Materials and Methods

### 2.1. Cases

From January 2008 to December 2017, patients diagnosed with adenomyosis and started conservative treatment at Kindai University Hospital and Osaka Red Cross Hospital were selected and studied retrospectively. Exclusion criteria was the absence of symptoms due to adenomyosis (e.g., if the patient is being monitored for endometriosis or other comorbidities) or pre-treatment imaging, request for hysterectomy at the first visit, presence of submucosal myoma or 3 cm/three or more intramuscular myomas that may cause hypermenorrhea.

### 2.2. Diagnosis

The diagnosis of adenomyosis was made using patients’ symptoms, such as dysmenorrhea and hypermenorrhea, and imaging techniques, such as MRI or transvaginal ultrasound. The criteria for diagnosis by MRI were the presence of an enlarged myometrium with an indistinct limbus and a heterogeneous internal signal on T2-weighted images or thickening of the junctional zone (>12 mm).^15 16^ The diagnostic criteria for transvaginal ultrasonography were asymmetrical enlargement of the myometrium and an asymmetrical decrease in echogenicity of the lesion.^1 10^ Most of the cases were diagnosed by MRI, but only two cases were diagnosed by transvaginal ultrasonography without pre-treatment MRI. Age was defined as the time when adenomyosis was diagnosed on imaging.

### 2.3. Size Measurement

Measurements of the size of the uterus and the myometrium were performed using MRI (figure1). In sagittal sections of MRI T2-weighted images, the length from the cervix to the bottom of the uterus was defined as the long axis diameter of the uterus (a), the maximum diameter perpendicular to long axis diameter was defined as the short axis diameter of the uterus (b), and the thickness of the uterine muscle layer within the short axis diameter of the uterus was defined as the muscle layer thickness (c). The maximum transverse diameter of the uterus in the axial section of MRI T2-weighted images was defined as the transverse diameter of the uterus (d). In the two cases measured by transvaginal ultrasonography, (a), (b), and (c) were measured at the position of maximum sagittal section.

**Figure 1.**
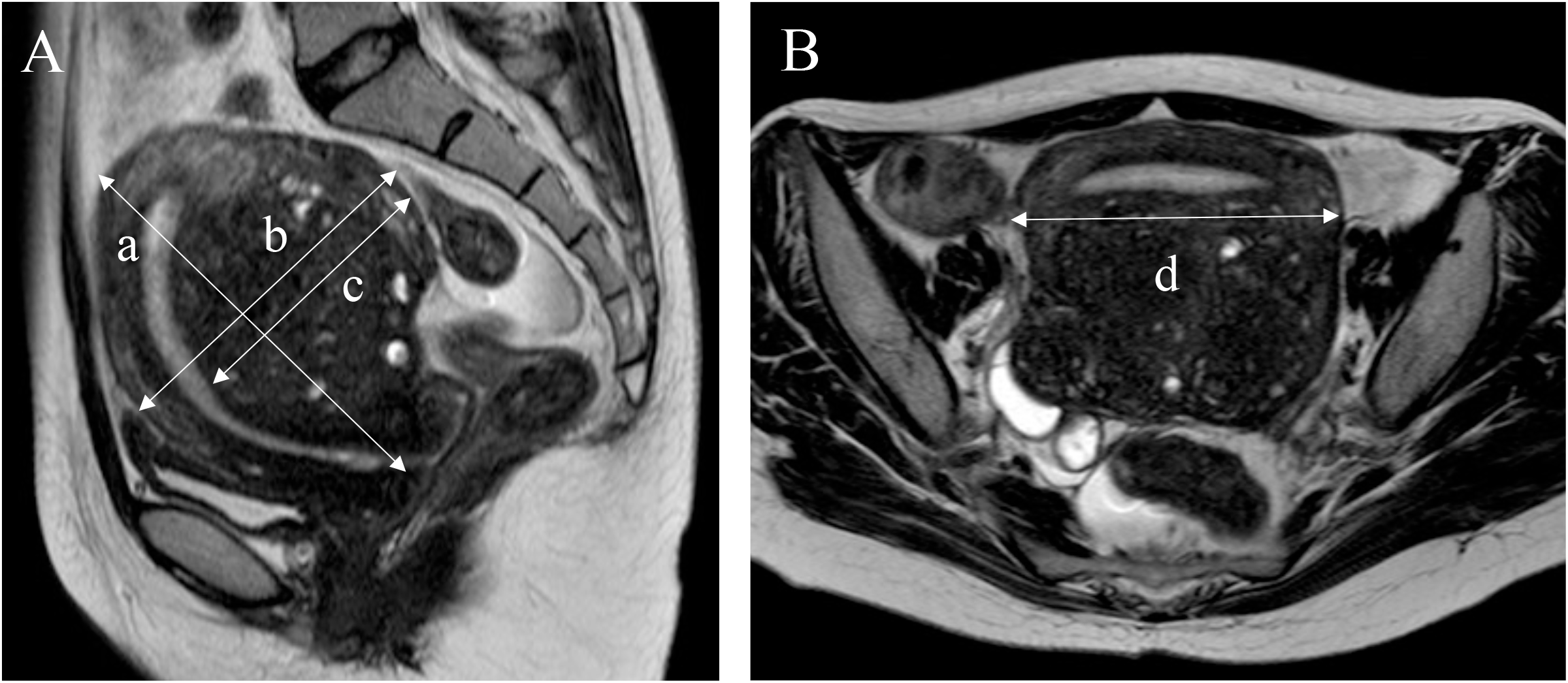
Measurement of uterine size. T2-Weighted Image (T2WI) of MRI. A: We used the sagittal T2WI of the uterus to measure (a); the uterine long axis diameter, (b); the uterine short axis diameter and (c); the muscle layer thickness. B: We used the axial T2WI of the uterus to measure (d); the uterine transverse diameter.

### 2.4. Type of Adenomyosis

Adenomyosis was classified into four subtypes based on MRI imaging features.^17^ Subtype I adenomyosis involved adenomyotic lesions that extended from the endometrium and did not extend to the entire myometrium. Subtype II adenomyosis was defined as adenomyotic lesions that extended from the perimetrium and did not extend into the junctional zone. Subtype III adenomyosis was an isolated adenomyotic lesion in the myometrium that did not extend into the junctional zone and the perimetrium. Subtype IV adenomyosis was defined as a lesion that could not be classified as types I–III, where the lesion involved the entire muscle layer. Two cases diagnosed by transvaginal ultrasonography were not evaluated.

### 2.5. Type of conservative treatment

Hormone therapy (gonadotropin releasing hormone agonist (GnRHa), progestins, levonorgestrel-releasing intrauterine system (LNG-IUS, Mirena intrauterine delivery system^®^, Bayer Yakuhin, Ltd), oral contraceptives (OCs), and danazol (BONZOL tablets^®^, Mitsubishi Tanabe Pharma Corporation) and adenomyomectomy were provided as conservative treatment for adenomyosis. Hysterectomy was performed after consultation with the patient when the symptoms worsened, or it became difficult to continue hormone therapy. Treatment was started on the date of the first visit, and the end of treatment was set at the date of the hysterectomy surgery or at the end of the observation period.

### 2.6. Statistical analysis

Statistical analysis was performed using Graphpad Prism ver. 8.2.0 (GraphPad Software, San Diego, CA, USA). The cumulative hysterectomy rate was determined by the Log-rank test, and comparison between the two groups used the Mann-Whitney U test and χ^2^ test, with P<0.05 as a significant difference. Decision tree analysis was performed using weka (https://doi.org/10.1016/j.knosys.2019.04.013).

### 2.7. Ethics statement

This study was conducted with the approval of the ethics committees of Kindai University Hospital and Osaka Red Cross Hospital (The approval numbers are R02-090 for Kindai University Hospital and J-0156 for Osaka Red Cross Hospital).

### 2.8. Patients and public involvement

Patients and the public were not involved in this study, including data collection, analysis and interpretation.

## 3. Results

A total of 885 patients were diagnosed with adenomyosis and started on treatment; 694 with no symptoms or no pre-treatment imaging, 51 who requested a hysterectomy at the time of first visit, and 16 with submucosal myoma or 3 cm/three or more intramuscular myomas that may cause hypermenorrhea were excluded, and 124 patients were started on conservative treatment (figure 2). Baseline characteristics of the 124 patients are presented in table 1. The median treatment period was 28 months (1– 132 months), median age was 41 years (24–53 years), median parity was 1 (0–3), median long axis diameter of the uterus was 9.7 cm (6.3–17.7 cm), median short axis diameter was 6.7 cm (3.5–12.9 cm), median transverse diameter was 6.8 cm (2.8–14.2 cm) and the median muscle layer thickness was 3.9 cm (1.3–8.8 cm). Adenomyosis subtypes I, II, III and IV were identified in 33 (26.6%), 28 (22.6%), 3 (2.4%) and 60 (48.4%) of these patients, respectively. Conservative treatment with hormone therapy alone was provided for 117 patients (94.4%), adenomyomectomy alone was performed for three patients (2.4%), and a mixture of these two procedures were provided for four patients (3.2%). The breakdown of hormone therapy is presented in figure 3.

**Table 1.** Clinical characteristics of the 124 patients undergoing conservative treatment for adenomyosis

|  | n=124 |
| --- | --- |
| Age | 41 (24-53) |
| Parity | 1 (0-3) |
| Gravida | 1 (0-6) |
| Size of Uterus |  |
| Long axis diameter (cm) | 9.65 (63-177) |
| Short axis diameter (cm) | 6.65 (35-129) |
| Transvers diameter (cm) | 6.75 (28-142) |
| Muscle layer thickness (cm) | 3.9 (13-88) |
| Type of Adenomyosis |  |
| Type I | 33 (26.6%) |
| Type II | 28 (22.6%) |
| Type III | 3 (2.4%) |
| Type IV | 60 (48.4%) |
| Type of Treatment |  |
| Hormonal Therapy | 117 (94.4%) |
| Adenomyomectomy | 3 (2.4%) |
| Hormonal & Adenomyomectomy | 4 (3.2%) |

**Figure 2.**
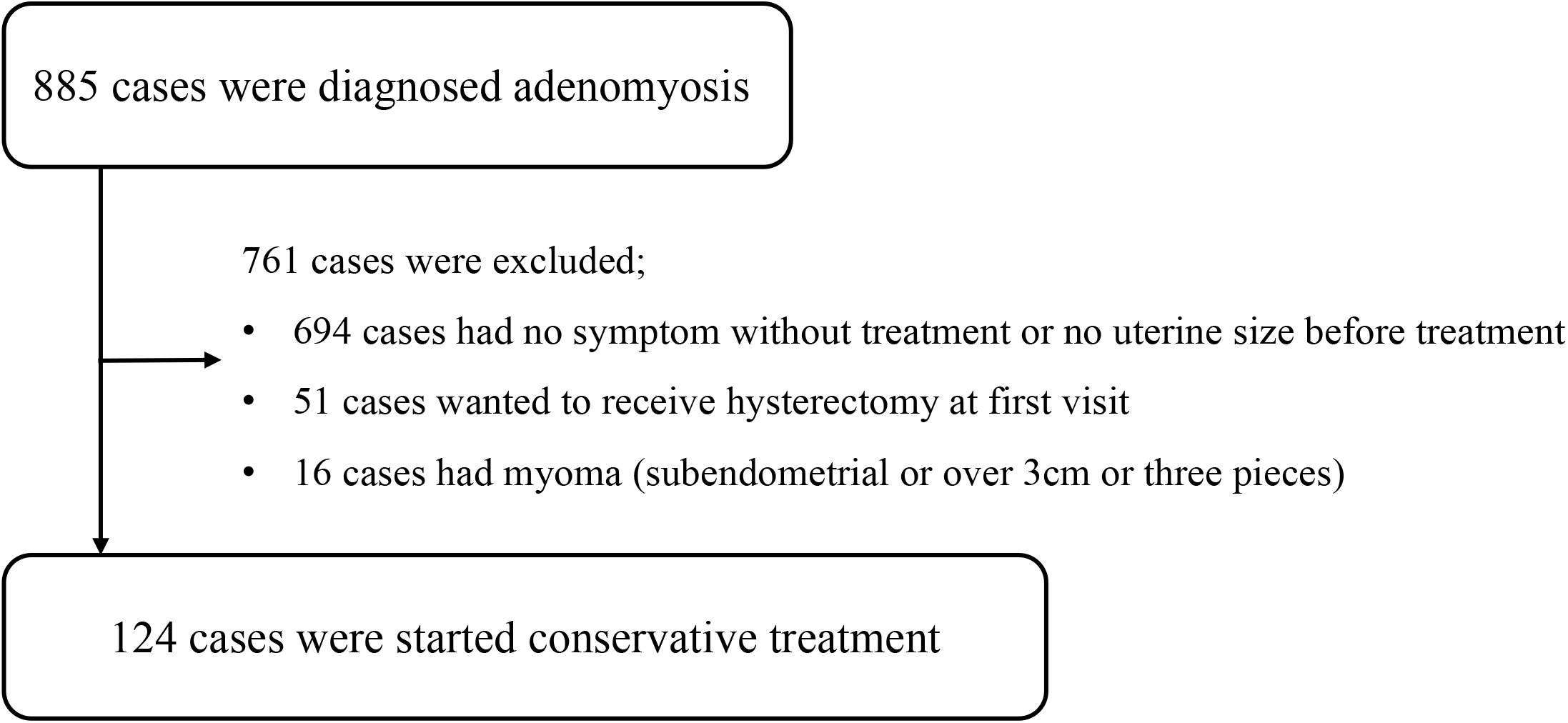
Cases flow chart. Of the 885 patients diagnosed with uterine adenomyosis, conservative treatment was initiated in 124 patients.

**Figure 3.**
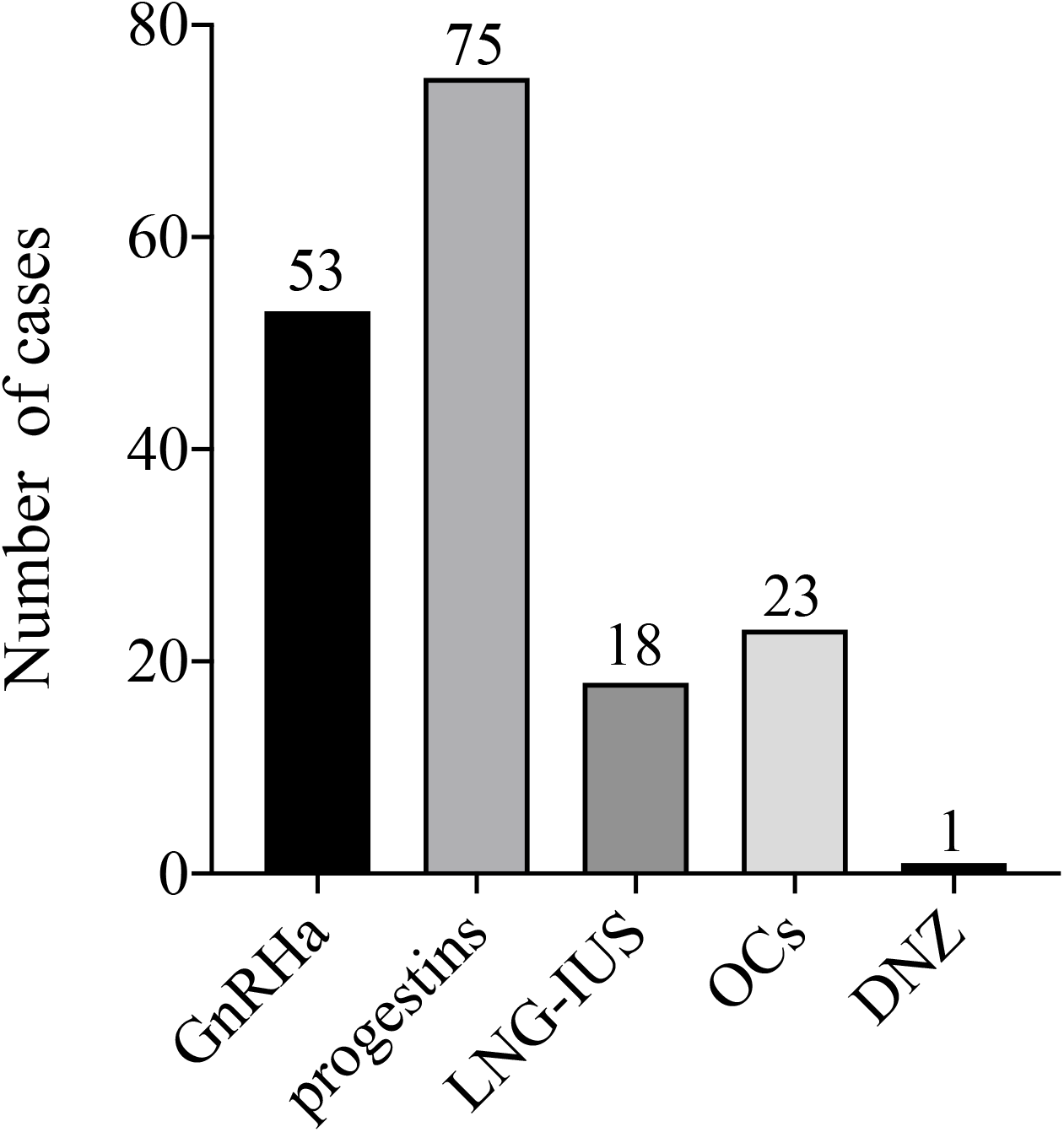
Number of cases treated with hormone therapy. GnRHa: gonadotropin releasing hormone agonist, LNG-IUS: levonorgestrel-releasing intrauterine systems, OCs: Oral contraceptives, DNZ: danazol. Y-axis shows the number of cases.

Ninety-six women (77.4%) were able to continue conservative treatment throughout the treatment period, and 28 patients (22.6%) required hysterectomy during the course of conservative treatment (figure 4). The cumulative total hysterectomy rate, determined from the log-rank test of 124 patients who started conservative treatment, was 32.4% and the 28 that required hysterectomy (Group A) all had hysterectomy within 63 months (figure 4). Of the 96 patients who were able to continue conservative treatment, 26 were able to continue conservative treatment for adenomyosis beyond 63 months (Group B), and all of them ultimately did not require hysterectomy (figure 4).

**Figure 4.**
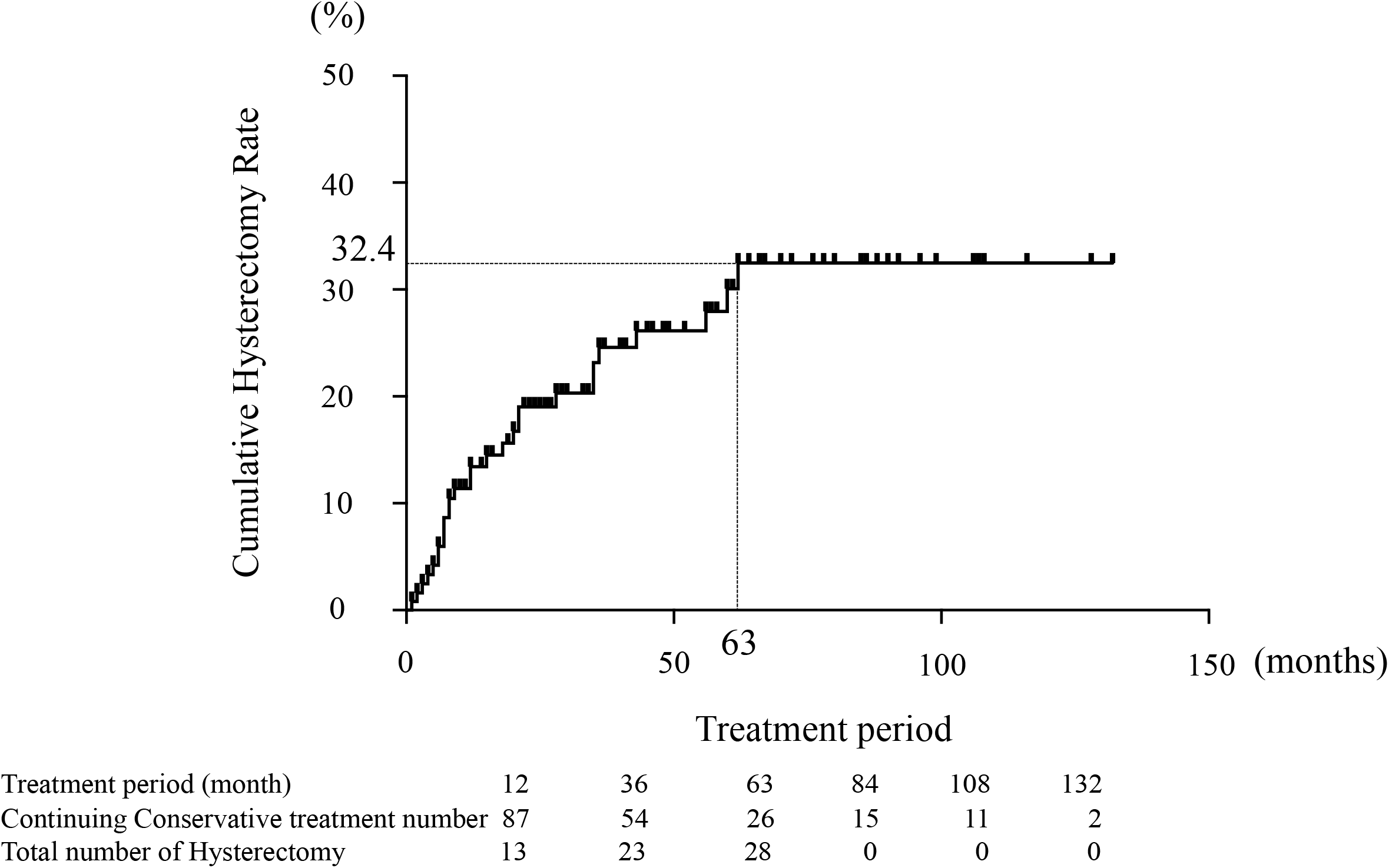
Cumulative hysterectomy rate. Kaplan-Meier analysis of the treatment period. The X-axis is the duration of treatment and Y-axis is the cumulative hysterectomy rate. The cumulative hysterectomy rate was 32.4% and reached a plateau after 63 months. The median treatment period was 28 months (1–132 months).

The characteristics of Group A and Group B are presented in Table 2. Group A had a significantly higher age (Group A: 43 years, Group B: 37 years, p<0.001), higher gravidity (Group A: 2, Group B: 0, p<0.001) and parity (Group A: 2, Group B: 0, p<0.001), and a significantly higher proportion of multipara (Group A: 82.1%, Group B: 42.3%, p<0.001) compared with Group B. The long axis diameter (Group A: 11.1 cm, Group B: 9.0 cm, p<0.001), short axis diameter (Group A: 9.0 cm, Group B: 7.7 cm, p=0.002), transverse diameter Group A: 8.0 cm, Group B: 6.6 cm, p=0.012), and muscle layer thickness (Group A: 4.6 cm, Group B: 3.6 cm, p=0.018) were significantly larger in Group A than those in Group B. The proportion of subtype IV adenomyosis and other complications of endometriosis were not significantly different between the two groups.

**Table 2.** Comparison of clinical characteristic of at baseline between cases failed conservative therapy (Group A) and continued uterine conservative therapy (Group B).

|  | Group A<br>(n=28) | Group B<br>(n=26) | P value |
| --- | --- | --- | --- |
| Age† | 43 (33-53) | 37 (27-46) | <0.001 |
| Gravida† | 2 (0-6) | 0 (0-3) | <0.001 |
| Parity† | 2 (0-3) | 0 (0-2) | <0.001 |
| Multipara† | 23 (82.1%) | 11 (42.3%) | <0.001 |
| Long axis diameter (cm)* | 11.1 (76-177) | 9.0 (64-130) | <0.001 |
| Short axis diameter (cm)* | 7.7 (47-129) | 6.0 (35-99) | 0.002 |
| Transvers diameter (cm)* | 8.0 (41-142) | 6.6 (37-92) | 0.012 |
| Muscle layer thickness (cm)* | 4.6 (25-73) | 3.5 (13-64) | 0.018 |
| Type IV adenomyosis* | 18 (64.3%) | 10 (38.5%) | 0.059 |
| Another of endometriosis* | 11 (39.3%) | 13 (50.0%) | 0.766 |

\*the Mann-Whitney U test, † $\chi^2$ test

To determine the critical factors involved in whether conservative treatment for symptomatic adenomyosis can be continued or not, we performed a decision tree analysis of Groups A (group of discontinued conservative treatment) and B (group of continued conservative treatment) using all the factors presented in Table 2, as shown in figure 5. Interestingly, only parity and age, rather than factors related to adenomyotic lesions, such as uterine size or adenomyosis subtype classification, had a decisive impact on the success rate of conservation treatment. The first and most important factor was parity, with 74% (23/31) of women with a parity of zero or one continuing conservative treatment, compared to only 13% (3/23) with a parity of two or more continuing conservative treatment. A total of 80% of patients were divided into two groups based on whether or not they could continue treatment with parity alone. For example, three cases of hysterectomy occurred in patients aged 47 years and older who had a parity of zero or one. When parity was two or more, only two patients younger than 38 years continued conservative treatment.

**Figure 5.**
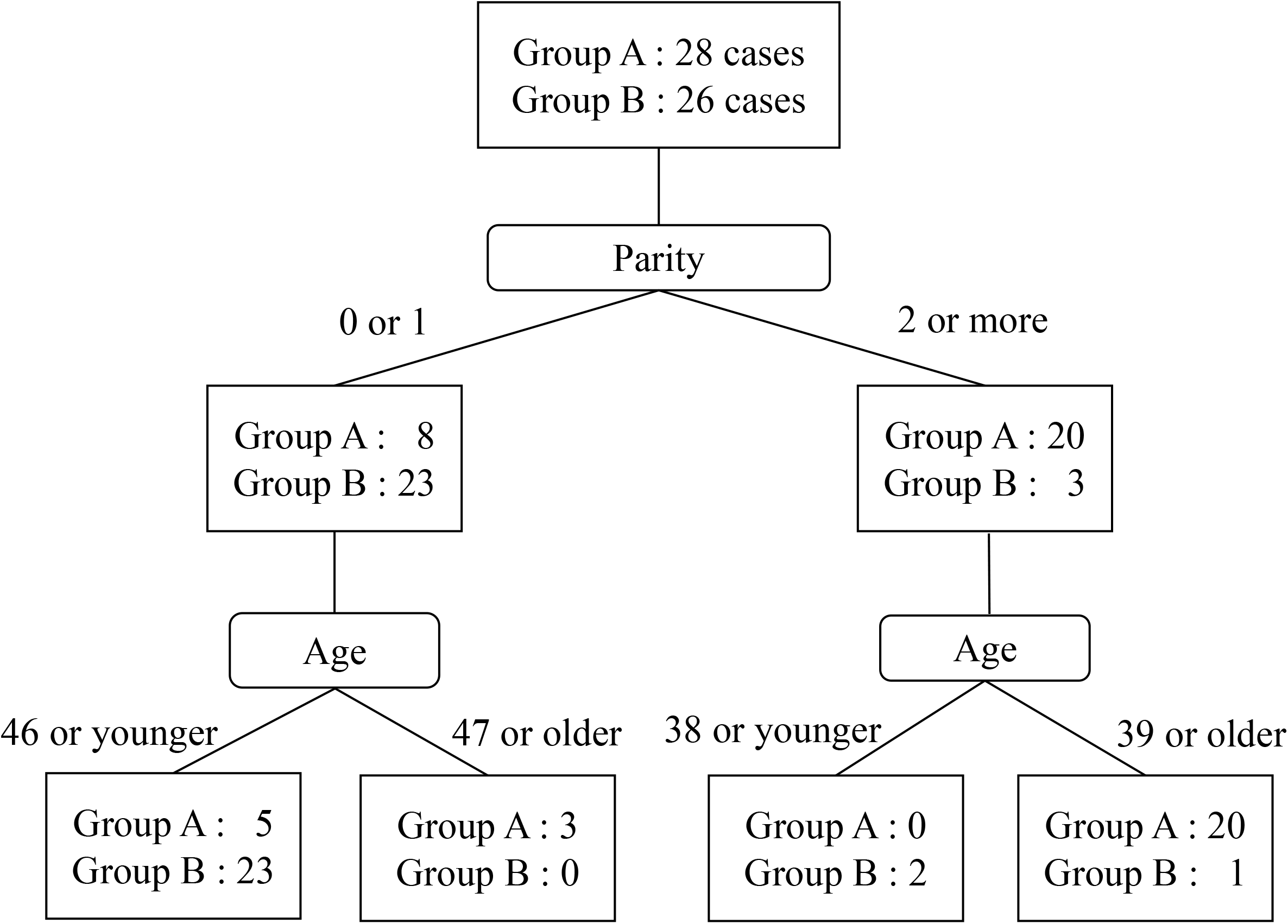
Decision tree analysis. Group A; cases that required hysterectomy, Group B; cases that continued conservative treatment of adenomyosis. Accuracy: 77.8%

## 4. Discussion

We retrospectively examined the course of attempted uterine preservation in patients with symptomatic adenomyosis to determine in which patient conservative treatment could be continued and in which patient hysterectomy was necessary. This study was unique in that (i) we extracted continued and discontinued conservative treatment cases from the curves of the cumulative hysterectomy rate increase in women who attempted uterine preservation, and (ii) we identified factors that distinguish between uterine preservation and non-preservation were clarified by decision tree analysis. For the first time, this study showed that parity and age may be important factors for the consideration of conservative treatment for adenomyosis. Women in this study were relatively young, with a median age of 41 years, making them younger than those in reports from the early 2000s, but more consistent with recent reports.^18-21^ The median parity was also low (at one), which may reflect the recent increase in aging of primipara and the trend of low fertility (United nations: World Population Prospects 2019). In previous reports examining the benefit of hormone therapy in adenomyosis, the mean pre-treatment uterine volume was 86 cm^3 22^, 96.5 cm^3 3^, 113.8 cm^3 23^, 158.9 ml ^24^, 278 cm^3 25^, and 311 cm^3 26^. The median uterine volume calculated from the long, short, and transverse uterine diameters in the present study was 217 (71–1400) cm^3^, so the size of the uterus was consistent with those previously reported. Adenomyosis was classified as subtype IV in half of the cases, which tended to be more severe than previously reported.^17^ This may be due to the fact that the two centers participating in the study were core hospitals in the region, and therefore accepting patients with advanced or difficult diagnosis. In conservative treatment for adenomyosis, adenomyomectomy is indicated when hormone therapy is difficult to continue or when the patient is undergoing infertility treatment. Because the uterine myometrium must be repaired after removal of the adenomyotic lesion, it is limited to lesions that are localized and capable of preserving the normal muscle layer ^27^. In the present study, adenomyomectomy was chosen for a very small number of cases during infertility treatment or when there was a desire for surgery. Multiple methods of hormone therapy were used in most cases, including GnRHa, OCs, progestins, LNG-IUS and danazol.

Multiple reports have shown that the smaller the size of the uterus at the start of hormone therapy, the more successful hormone therapy has been in treating adenomyosis ^3 22-25 28 29^. In this study, the size of the uterus at the start of treatment was also significantly smaller in Group B, which was able to continue with conservative treatment of adenomyosis (table 1). However, previous reports have had mixed follow-up periods and may have included women who ultimately needed hysterectomy. In the present study, of the 124 patients who started conservative treatment, the failure to continue treatment and necessary hysterectomy were most frequent within the first year. This frequency then decreased, and treatment continued in all patients without much change until the fifth year. Women who were able to continue conservative treatment beyond 63 months did not require a hysterectomy. This novel analysis and the above results may provide guidance for planning the treatment of adenomyosis. Furthermore, our study exploring factors involved in the acceptability of conservative treatment found that patients undergoing conservative treatment for at least 5 years should be compared with those who have had hysterectomy.

Decision tree analysis was able to extract the fewest factors needed to most clearly separate the two patient groups (Group A and B) in terms of sensitivity and specificity. Surprisingly, our current study revealed that uterine size and adenomyosis subtype classification^17^ were not among the factors. The most important factor was parity, and most patients with a parity of two or more were found to eventually require hysterectomy. This may reflect the psychological factor of patients with two or more children wanting to prioritize parenthood, rather than continuing conservative treatment, which is also associated with symptoms such as irregular bleeding and pain. It has been reported that patients who had undergone hysterectomy for any condition, not just adenomyosis, were significantly more likely to have had a parity of two or more.^4^ In addition, parity was reported to correlate with the incidence of adenomyosis^11 30^, which may have influenced this result. The second most important factor in the decision tree analysis was age. This may also be related to the intensity of the desire to preserve the uterus and the frequency of adenomyosis. Parity and age of diagnosis may be stratifying factor in future clinical trials on hormone therapy.

One limitation of this study was the small number of cases. We screened 885 cases of adenomyosis, but only 124 patients matched the criteria for inclusion in the analysis. The number of cases was further reduced to 26, because we found that only patients successfully treated for more than 63 months could be considered to have successful uterine preservation. Therefore, it is expected that about 5000 patients with adenomyosis would be needed to perform a similar analysis with more than 100 cases per group. Furthermore, this study was a retrospective study of routine practice over a 10-year period, and the diversity in treatments available over this period is also a limitation. In Japan, progestins and the levonorgestrel-releasing intrauterine system were approved within the last 5 years for the treatment of adenomyosis, the increased frequency of their use may have influenced the results. Additionally, criteria for the diagnosis of adenomyosis are not yet clear, so it is possible that the patient may not be diagnosed even if she has symptoms. It is hoped that further research will establish clear criteria for the diagnosis and treatment of adenomyosis.

## 5. Conclusions

Uterine preservation in patients with adenomyosis is more likely to be successful if they can continue conservative treatment for approximately 5 years. In addition, multipara and higher age at diagnosis are factors for hysterectomy during conservative treatment of adenomyosis. The results of this study may be useful in decision-making and for informed consent when treating patients with adenomyosis. Parity and age at diagnosis may be stratifying factor in future clinical trials on hormone therapy.

## Data Availability

We replaced special formatting or symbols to the appropriate codes.

## Notes

### Competing Interest Statement

The authors have declared no competing interest.

### Funding Statement

No external funding was received

### Author Declarations

The ethics committees of Kindai University Hospital and Osaka Red Cross Hospital (The approval numbers are R02-090 for Kindai University Hospital and J-0156 for Osaka Red Cross Hospital).

## References

1. Levy G, Dehaene A, Laurent N, et al. An update on adenomyosis. Diagn Interv Imaging. 2013;94(1):3–25.

2. Vannuccini S, Luisi S, Tosti C, Sorbi F, Petraglia F. Role of medical therapy in the management of uterine adenomyosis. Fertil Steril. 2018;109(3):398–405.

3. Osuga Y, Fujimoto-Okabe H, Hagino A. Evaluation of the efficacy and safety of dienogest in the treatment of painful symptoms in patients with adenomyosis: a randomized, double-blind, multicenter, placebo-controlled study. Fertil Steril. 2017;108(4):673–678.

4. Vercellini P, Parazzini F, Oldani S, Panazza S, Bramante T, Crosignani PG. Adenomyosis at hysterectomy: a study on frequency distribution and patient characteristics. Hum Reprod. 1995;10(5):1160–1162.

5. Parazzini F, Mais V, Cipriani S, Busacca M, Venturini P, On behalf of GISE. Determinants of adenomyosis in women who underwent hysterectomy for benign gynecological conditions: results from a prospective multicentric study in Italy. Eur J Obstet Gynecol Reprod Biol. 2009;143(2):103–106.

6. Ryan GL, Stolpen A, Van Voorhis BJ. An unusual cause of adolescent dysmenorrhea. Obstet Gynecol. 2006;108(4):1017–1022.

7. Farquhar C, Brosens I. Medical and surgical management of adenomyosis. Best Pract Res Clin Obstet Gynaecol. 2006;20(4):603–616.

8. Dueholm M, Lundorf E. Transvaginal ultrasound or MRI for diagnosis of adenomyosis. Curr Opin Obstet Gynecol. 2007;19(6):505–512.

9. Pinzauti S, Lazzeri L, Tosti C, et al. Transvaginal sonographic features of diffuse adenomyosis in 18-30-year-old nulligravid women without endometriosis: association with symptoms. Ultrasound Obstet Gynecol. 2015;46(6):730–736.

10. Garcia L, Isaacson K. Adenomyosis: review of the literature. J Minim Invasive Gynecol. 2011;18(4):428–437.

11. Vannuccini S, Petraglia F. Recent advances in understanding and managing adenomyosis. F1000Res. 2019;8.

12. Struble J, Reid S, Bedaiwy MA. Adenomyosis: A Clinical Review of a Challenging Gynecologic Condition. J Minim Invasive Gynecol. 2016;23(2):164–185.

13. Benetti-Pinto CL, Mira TAA, Yela DA, Teatin-Juliato CR, Brito LGO. Pharmacological Treatment for Symptomatic Adenomyosis: A Systematic Review. Rev Bras Ginecol Obstet. 2019;41(9):564–574.

14. Younes G, Tulandi T. Conservative Surgery for Adenomyosis and Results: A Systematic Review. J Minim Invasive Gynecol. 2018;25(2):265–276.

15. Dueholm M, Lundorf E, Hansen ES, Sørensen JS, Ledertoug S, Olesen F. Magnetic resonance imaging and transvaginal ultrasonography for the diagnosis of adenomyosis. Fertil Steril. 2001;76(3):588–594.

16. Novellas S, Chassang M, Delotte J, et al. MRI characteristics of the uterine junctional zone: from normal to the diagnosis of adenomyosis. AJR Am J Roentgenol. 2011;196(5):1206–1213.

17. Kishi Y, Suginami H, Kuramori R, Yabuta M, Suginami R, Taniguchi F. Four subtypes of adenomyosis assessed by magnetic resonance imaging and their specification. Am J Obstet Gynecol. 2012;207(2):114.e111-117.

18. Levgur M, Abadi MA, Tucker A. Adenomyosis: symptoms, histology, and pregnancy terminations. Obstet Gynecol. 2000;95(5):688–691.

19. Templeman C, Marshall SF, Ursin G, et al. Adenomyosis and endometriosis in the California Teachers Study. Fertil Steril. 2008;90(2):415–424.

20. Taran FA, Weaver AL, Coddington CC, Stewart EA. Understanding adenomyosis: a case control study. Fertil Steril. 2010;94(4):1223–1228.

21. Pontis A, D’Alterio MN, Pirarba S, de Angelis C, Tinelli R, Angioni S. Adenomyosis: a systematic review of medical treatment. Gynecol Endocrinol. 2016;32(9):696–700.

22. Li L, Leng J, Jia S, Lang J. Treatment of symptomatic adenomyosis with the levonorgestrel-releasing intrauterine system. Int J Gynaecol Obstet. 2019;146(3):357–363.

23. Sheng J, Zhang WY, Zhang JP, Lu D. The LNG-IUS study on adenomyosis: a 3-year follow-up study on the efficacy and side effects of the use of levonorgestrel intrauterine system for the treatment of dysmenorrhea associated with adenomyosis. Contraception. 2009;79(3):189–193.

24. Lee KH, Kim JK, Lee MA, et al. Relationship between uterine volume and discontinuation of treatment with levonorgestrel-releasing intrauterine devices in patients with adenomyosis. Arch Gynecol Obstet. 2016;294(3):561–566.

25. Fawzy M, Mesbah Y. Comparison of dienogest versus triptorelin acetate in premenopausal women with adenomyosis: a prospective clinical trial. Arch Gynecol Obstet. 2015;292(6):1267–1271.

26. Zhang P, Song K, Li L, Yukuwa K, Kong B. Efficacy of combined levonorgestrel-releasing intrauterine system with gonadotropin-releasing hormone analog for the treatment of adenomyosis. Med Princ Pract. 2013;22(5):480–483.

27. Abbas S, Raybould JE, Sastry S, de la Cruz O. Respiratory viruses in transplant recipients: more than just a cold. Clinical syndromes and infection prevention principles. Int J Infect Dis. 2017;62:86–93.

28. Shaaban OM, Ali MK, Sabra AM, Abd El Aal DE. Levonorgestrel-releasing intrauterine system versus a low-dose combined oral contraceptive for treatment of adenomyotic uteri: a randomized clinical trial. Contraception. 2015;92(4):301–307.

29. Neriishi K, Hirata T, Fukuda S, et al. Long-term dienogest administration in patients with symptomatic adenomyosis. J Obstet Gynaecol Res. 2018;44(8):1439–1444.

30. Vercellini P, Viganò P, Somigliana E, Daguati R, Abbiati A, Fedele L. Adenomyosis: epidemiological factors. Best Pract Res Clin Obstet Gynaecol. 2006;20(4):465–477.

